# Antibodies against influenza A/H1N1pdm2009 and B/Victoria strains but not A/H3N2 are increased in recent onset type 1 narcolepsy versus matched controls

**DOI:** 10.64898/2026.06.13.26355596

**Authors:** Han Yan, Ling Lin, Robin Guillard, Jing Zhang, Claudia Macaubas, Fabio Pizza, Francesco Biscarini, Giuseppe Plazzi, Vamsee Mallajosyula, Mark Davis, Holden Maecker, Emmanuel Mignot

## Abstract

**Study Objectives:** Onsets of Narcolepsy type-1 (NT1) increased following A/H1N1 vaccination with Pandemrix® in Europe and with A/H1N1pdm2009 infections in China and other countries. To test if other strains could trigger narcolepsy, we measured strain-specific antibodies in patients with recent onset NT1 compared to controls.

**Methods:** Antibodies against hemagglutinin (HA) and neuraminidase (NA) were tested in 62 patients with very recent onset (onset and blood collection following a single flu season, mean ± SEM: 0.44 ± 0.06 years since onset) and 100 controls matched by age, sex, season and year of collection (2000-2025). Results were next extended to 181 recent onset patients (mean± SEM: 1.00 ± 0.05 years) versus 260 controls, matched by sex, season and year, but having a slightly higher mean age. HA inhibition (HAI) and NA inhibition (NAI) assays were conducted using flu strains known to circulate during the corresponding flu seasons. HAI results are shown as % positive (titers ≥ 40) and NAI results as geometric mean titers. Odds ratio (OR) and β coefficient were used to compare antibody titers in NT1 versus controls. The contribution of each assay to prediction was finally quantified in the larger sample set using Shapley decomposition.

**Results:** NT1 patients had increased anti-HA and anti-NA antibodies against A/H1N1pdm2009 (anti-HA OR= 3.86, anti-NA β= 0.35) and B/Victoria (anti-HA OR=1.90, anti-NA β=0.22), but not A/H1N1pre2009, A/H3N2, or B/Yamagata, independent of HLA-DQB1*06:02 status, age, sex, and flu season. Correlations between anti-HA and anti-NA antibodies titers were weak to moderate but significant (r^2^=-0.10 to 0.34). Multivariable model outperformed age-only baseline (McFadden R^2^ = 0.19 vs. 0.03; AUC = 0.79 vs. 0.64; likelihood-ratio test χ^2^ = 51, p<10^-9^), with anti-HA against A/H1N1pdm2009 (β = 0.78, p < 10^-6^) and anti-NA against B/Victoria (β = 0.69, p < 10^-5^) emerging as the strongest independent predictors.

**Conclusions:** A/H1N1pdm2009 and B/Victoria, but not other strains can trigger the autoimmune process leading to orexin cell loss in narcolepsy.

## Introduction

Approximately 0.03% of the population across ethnic groups and in various countries are affected by narcolepsy-cataplexy, also known as narcolepsy type 1 (NT1)^1^. This disorder is highly associated with a unique human leukocyte antigen (HLA) heterodimer, DQA1*01:02/DQB1*06:02, and a loss of ∼ 70,000 hypothalamic neurons producing the wake promoting peptide orexin (also called hypocretin)^2,3^. Additional genetic associations are identified through genome-wide association studies include other HLA effects, T-cell receptor (TCR) loci, other autoimmune disease loci such as TNFSF4, CTSH, ZFN365, SIRPG, or genes involved in immune responses to allergy and infection (langerin and INFAR1)^4^, supporting autoimmunity as the likely cause of orexin deficiency in narcolepsy.

Despite strong indirect genetic evidence, definitive proof that autoimmunity mediates orexin cell loss in narcolepsy, for example presence of autoantibodies, is still lacking^2^. Increased T-cell responses directed toward orexin have been found by multiple investigators^5–7^ especially in recent onset patients, but these responses are not specific and occur at lower rate in controls. Further, T-cell phenotypes responsive to orexin peptides do not differ in narcolepsy patients vs controls^5,7^.

More convincingly, onset of narcolepsy has been linked with pandemic H1N1 2009 Influenza-A following infections^8,9^ and vaccinations (using Pandemrix®)^10^ in US^8^, China9 and Europe^10–12^ through epidemiological studies respectively. More recently, another peak of increased incidence of narcolepsy was observed in Europe in 2013^13^, without any clear association with A/H1N1. Rather, a suggestion of association with influenza B was made based on abundance of this infection in the 2012-2013 winter season in Europe. In this work, we tested whether recent onset patients with NT1 have increased antibodies titers against various flu stains, comparing patients to carefully matched controls, as flu infection type varies by year and age1^4,15^.

## Methods

### Subjects

Blood samples were collected from 2000 to 2025, and sera stored at −80 ℃ until use. All patients met International Classification of Sleep Disorders 3 (ICSD-3) criteria for NT1. The study was approved by an Institutional Review Board of Stanford University, and all participants gave written informed consent or assent (if applicable). A primary sample set of 62 recent-onset NT1 cases (56.5% male; mean age ± standard error of the mean [SEM] 11.2 ± 1.11 [3 - 48] years; mean age of onset ± SEM 10.7 ± 1.13 [2.7 - 48] years; mean interval between narcolepsy onset and serum collection ± SEM 0.44 ± 0.06 [0 - 1] years) was selected based on the criteria that only one flu season had preceded disease onset and blood collection. These were carefully matched with samples from 100 unrelated controls (deliberately enriched in DQB1*06:02 positive controls) with similar age (mean ± SEM 12.6 ± 0.39[3-21] years), sex (46% male), and flu season when collected; a slightly larger share of controls was white (Table 1). Cerebrospinal fluid (CSF) hypocretin-1 levels were available for 29 patients, and all lower than 110 pg/ml (mean ± SEM 22.7 ± 4.68 pg/ml).

**Table 1.** Demographic, HLA status and sample characteristics of the very recent onset NT1 patients and controls.

|  | Patients<br>(N=62) | DQ0602+<br>Controls<br>(N=55) | DQ0602-<br>Controls<br>(N=45) | Total<br>Controls<br>(N=100) | p-value |
| --- | --- | --- | --- | --- | --- |
| Age (mean ± SEM<br>[yrs] [N]) | 11.2 ± 1.11<br>(62) | 13.0 ± 0.51<br>(55) | 12.1 ± 0.60<br>(45) | 12.6 ± 0.39<br>(100) | 0.16 |
| Sex (N [M/F]) | 35/27 | 21/34 | 25/20 | 46/54 | 0.20 |
| Interval* (mean ± SEM<br>[yrs] [N]) | 0.44 ± 0.06<br>(62) | N/A | N/A | N/A | N/A |
| HLA-DQB1*0602<br>(%[N]) | 100% (62) | 100% (55) | 0% (45) | 55% (100) | N/A |
| CSF Orexin (mean ±<br>SEM [pg/mL] [N]) | 22.7 ± 4.68<br>(29) | 323.2 ± 14.9<br>(2) | 327.8 ± 8.60<br>(6) | 326.7 ± 7.48<br>(8) | <0.001 |
| Season of sample<br>collection |  |  |  |  |  |
| Spring (N [%]) | 19 (30.6%) | 19 (34.5%) | 16 (35.6%) | 35 (35.0%) | 0.57 |
| Summer (N [%]) | 21 (33.9%) | 16(29.1%) | 8 (17.8%) | 24 (24.0%) | 0.17 |
| Autumn (N [%]) | 17 (27.4%) | 13 (23.6%) | 12 (26.7%) | 25(25.0%) | 0.73 |
| Winter (N [%]) | 5 (8.06%) | 7 (12.7%) | 9 (20.0%) | 16(16.0%) | 0.14 |
| White (N [%]) | 29/58<br>(50.0%) | 45/53<br>(84.9%) | 33/38<br>(86.8%) | 78/91<br>(85.7%) | <0.001 |

\* Defined as time duration from narcolepsy onset to serum collection.

The sample set was then extended to 181 patients with NT1 (51.4% male; mean age ± SEM 15.7 ± 0.86 [3 - 62] years; mean age of onset ± SEM 14.7 ± 0.85 [2.7 - 61] years, mean interval ± SEM 1.00 ± 0.05 [0 - 2] years) and 260 controls (46.5% male), which were overall older (mean age ± SEM 22.0 ± 0.76 [3 - 58] years, deliberately enriched in DQB1*06:02 positive, 50% each) (Table 2). CSF hypocretin-1 levels were available for 72 patients, showing low levels (<110 pg/ml) in all cases (mean ± SEM 20.1 ± 3.09 pg/ml).

**Table 2.** Demographic, HLA status and sample characteristics of the extended samples of NT1 patients and controls.

|  | Patients<br>(N=181) | DQ0602+<br>Controls<br>(N=154) | DQ0602-<br>Controls<br>(N=106) | Total<br>Controls<br>(N=260) | p-<br>value |
| --- | --- | --- | --- | --- | --- |
| Age (mean ± SEM<br>[yrs] [N]) | 15.7 ± 0.86<br>(181) | 23.3 ± 1.03<br>(154) | 20.2 ± 1.07<br>(106) | 22.0 ±<br>0.76(260) | <0.01 |
| Sex (N [M/F]) | 93/88 | 70/84 | 51/55 | 121/139 | 0.32 |
| Interval* (mean ±<br>SEM [yrs] [N]) | 1.00 ± 0.05<br>(181) | N/A | N/A | N/A | N/A |
| HLA-DQB1*0602<br>(%[N]) | 100% (181) | 100% (154) | 0% (106) | 59.2% (260) | N/A |
| CSF Orexin (mean<br>± SEM [pg/mL]<br>[N]) | 20.1 ± 3.09<br>(72) | 310.6 ± 22.8<br>(6) | 341.6 ± 10.3<br>(10) | 330.0 ±<br>101.3 (16) | <0.01 |
| Season of sample<br>collection |  |  |  |  |  |
| Spring (N [%]) | 62 (34.3%) | 49 (31.8%) | 28 (26.4%) | 77 (29.6%) | 0.30 |
| Summer (N [%]) | 48 (26.5%) | 49(31.8%) | 22 (20.7%) | 71 (27.3%) | 0.85 |
| Autumn (N [%]) | 35 (19.3%) | 36 (23.4%) | 35 (33.0%) | 71 (27.3%) | 0.05 |
| Winter (N [%]) | 36 (19.9%) | 20 (13.0%) | 21 (19.8%) | 41 (15.8%) | 0.26 |
| White (N [%]) | 136/181<br>(75.1%) | 137/154<br>(89.0%) | 92/106 (86.8<br>%) | 229/260<br>(88.1%) | <0.01 |

\* Defined as time duration from narcolepsy onset to serum collection.

### Hemagglutinin Inhibition (HAI) Assay

HAI assays were conducted following World Health Organization (WHO) protocol^16^. Briefly, amounts of hemagglutinin (HA) to be used for each strain were first determined by observing agglutination across serial 2-fold dilutions of virus when incubated with 1% Guinea pig red blood cells (gRBC). Subsequently, sera of patients and controls pretreated with Receptor Destroying Enzyme (RDE) are tested for inhibition of hemagglutination. This is done by testing serial 2-fold dilutions of sera from NT1 patients or controls starting from 1/10 dilution, incubating with 4 HA units for 0.5 hour, then adding 1% gRBC and incubating for another 1 hour. The highest dilution which inhibits agglutination is determined as the “effective’” antibody titer. Antibody titer ≥ 40 was used as a cut-off value to determine positivity in HAI assays, indicating infections within a year, and associated immune protection against the corresponding strain, for example in vaccination studies^17^.

### Neuraminidase Inhibition (NAI) Assay

NAI assays used the Enzyme-Linked Lectin Assay (ELLA) method^18^. In this assay, the terminal sialidase activity of recombinant neuraminidase (NA) is measured using fetuin as a substrate. Fetuin, a glycoprotein rich in sialic acid, is attached on a solid phase and NA activity reveals galactose residues. A Peroxidase-labeled peanut agglutinin (PNA) recognizing galactose is added next, and color revealed by measuring Optical Density (OD) at 450/650 nm following incubation with an enzyme substrate-chromogen reagent that measures unmasked galactose. The optimal strain-specific amount of NA used to assess inhibition by sera of NT1 patients or control is the amount that produces 90% of maximal activity on the assay. Sera pretreated with Receptor Destroying Enzyme (RDE) and serially diluted by 2-fold increments starting from a 1/10 dilution, are incubated first with the neuraminidase for 20 hours, then with PNA and its substrate-chromogen reagent. The titer of a tested serum is calculated as the reciprocal of the last dilution with an 450/650 OD equal to the cut-off value corresponding to a 50% end-point titer.

### Antigens used in HAI and recombinant NA used in NAI

Each sample was tested for HA and NA antibodies using HAI and NAI assays targeting the dominant strains known to have circulated at flu seasons of narcolepsy onset for patients, or at time of collection for controls. Dominant historical strains of A/H1N1, A/H3N2, B/Victoria, and B/Yamagata in North Hemisphere from 1999-2000 to 2024-2025 were selected based on WHO recommendations for influenza vaccine composition for the North Hemisphere^19^, and Center for Disease Control (CDC) Morbidity and Mortality Weekly Report (MMWR) of each Influenza season^20^.Strains used for HAI assay were purchased from Microbiologics, San Diego, CA, USA. Recombinant neuraminidases used in NAI assay were acquired from Sino Biological, Houston, Texas, US (Table S1).

### Statistical Analysis

Data was presented as mean ± SEM or %. Groups were primarily compared using Pearson χ2 or Student *t*-tests. Antibody titers to various influenza strains were grouped into A/H1N1pre2009 or A/H1N1pdm2009, A/H3N2, B/Victoria and B/Yamagata. A value of 5 was assigned as titer if test result was <10. Log10 transformed titers were then used for analysis. Geometric mean titers (GMT) were calculated as mean of log10 transformed titers. Titer ≥ 40^17^ was used as a well-known cutoff value for being positive for anti-HA antibodies. These are standard methods in influenza research.

Multivariate linear and logistic regressions were used to analyze the effect of NT1 on antibody titers after controlling for age, sex, DQB1*06:02, year and season. Covariates were included in the final model only when significant.

In addition, multivariate logistic regression model was performed to study association between narcolepsy diagnosis and a combination of anti-HA and anti-NA antibody variables as directed against A/H1N1pdm2009 and B/Victoria. To isolate the contribution of the predictors of interest beyond the confounder effect of age, we fitted two nested models on the same set of complete cases: a baseline model including age only (M₀: *narcolepsy diagnosis ∼ age*), and a full model (M₁: *narcolepsy diagnosis ∼ age + anti-HA against A/H1N1pdm2009 + anti-NA against A/H1N1pdm2009 + anti-HA against B/Victoria + anti-NA against B/Victoria*). Predictors were first preprocessed by applying base 10 logarithm to antibody titers and then standardized (z-scored) prior to model fitting to facilitate comparison of effect sizes. To assess global model performance, we used three complementary metrics. McFadden’s pseudo-R^2^ (1 − ℓ_full / ℓ_null, where ℓ denotes the log-likelihood) was reported as the standard likelihood-based fit index for logistic models. Tjur’s coefficient of discrimination, defined as the difference in mean predicted probabilities between cases and controls, was reported as a directly interpretable measure of class separation bounded in [0, 1]. The area under the receiver operating characteristic curve (AUC) is reported as a rank-based measure of discrimination. The incremental value of the predictors of interest over age alone was tested using a likelihood ratio test (M₁ vs. M₀), with 4 degrees of freedom corresponding to the number of added predictors.

To quantify the unique contribution of each predictor of interest, we decomposed each global performance metric (McFadden R^2^, Tjur D, AUC) using Shapley values. This method provides a principled attribution of explained performance and is robust to multicollinearity, overcoming limitations of leave-one-out ΔR^2^ approaches.

These analyses were performed in Python (version 3.12.10) using statsmodels for model fitting, scikit-learn for discrimination metrics, and custom code for the Shapley-based decomposition.

Pearson correlation analysis was performed between anti-HA and anti-NA antibodies titers against the same strain on the same samples.

## Results

### Recent onset narcolepsy is associated with increased titers of anti-HA and anti-NA antibodies directed against influenza 2009H1N1pdm and B/Victoria

The primary analysis of this study used samples of narcolepsy patients with very recent onset and blood collection unseparated by a flu season. Consequently, a unique flu season preceded both onset and blood sample collection. Controls matched for age, sex and flu season of collection were recruited concurrently across the same 25-year time window (2000 to 2025) (Table 1). In this carefully matched sample set, recent onset NT1 patients had increased percentage of samples positive for anti-HA antibodies against A/H1N1pdm2009 strain (anti-HA antibody titers ≥ 40) compared to controls (68.6% vs 44.8%), resulting in a 2.6 times increased risk of having been infected or vaccinated recently with A/H1N1pdm2009 compared to controls. Importantly, these patients also had increased percentage of samples positive for anti-HA antibodies against B/Victoria when compared to controls (33.3% vs 21.0%), with recent onset NT1 patients having a 2.13 times increased risk of past B/Victoria infections when compared to controls (Table 3).

**Table 3.** Hemagglutinin and neuraminidase antibody levels in recent onset NT1 patients and controls.

| Subtypes/lineages | Hemagglutinin Inhibition Assay (HAi) |  |  |  | Neuraminidase Inhibition Assay (NAI) |  |  |  |
| --- | --- | --- | --- | --- | --- | --- | --- | --- |
|  | % of positive* patients (N) | % of positive* controls (N) | Odds Ratio(95%CI) | p-value# | GMT (95CI) of patients (N) | GMT (95CI) of controls (N) | β-coefficient (95%CI) | p-value # |
| Influenza A |  |  |  |  |  |  |  |  |
| All H1N1 | 71.0% (62) | 55.0% (100) | 1.48(0.68,3.22) | 0.32 | 236.7(129.1,434.2) (45) | 124.2(83.5,184.6) (93) | 0.17(-0.10,0.45) | 0.21 |
| H1N1pre2009 | 73.3% (30) | 69.0% (42) | 1.11(0.38,3.25) | 0.86 | 33.6(19.8,57.2) (19) | 32.8(23.1,46.6) (42) | 0.01(-0.26,0.28) | 0.94 |
| H1N1pdm2009 | 68.8% (32) | 44.8% (58) | 3.60(1.11,11.6) | 0.03 | 1062.1(681.9,1654.2) (26) | 371.6(227.2,607.7) (51) | 0.51(0.20,0.82) | <0.01 |
| H3N2 | 40.3% (62) | 35.0% (100) | 1.20(0.62,2.32) | 0.59 | 47.9(29.9,76.9) (45) | 64.9(50.5,83.5) (93) | -0.18(-0.38,0.03) | 0.09 |
| Influenza B |  |  |  |  |  |  |  |  |
| B/Victoria | 33.3% (57) | 21.0% (100) | 3.13(1.86,10.2) | 0.01 | 445.8(286.1,694.5) (45) | 189.9(140.1,257.5) (93) | 0.29(0.03,0.54) | 0.03 |
| B/Yamagata | 14.0% (50) | 9.68% (93) | 1.49(0.52,4.26) | 0.46 | 236.7(177.7,315.4) (45) | 177.8(143.3,220.6) (92) | -0.08(-0.24,0.08) | 0.31 |

\*Using 40 as a cut-off value for being positive for HA antibody.
#Adjusted by age, sex, or other covariates when significant.

To confirm these results independently, we also conducted NAI assay. Using this assay, recent onset NT1 patients also had increased anti-NA antibody levels against A/H1N1pdm2009 (GMT: 1062.1 vs 371.6) and B/Victoria (GMT: 445.8 vs 189.9), with GMT of anti-NA antibodies against A/N1N1pdm2009 and B/Victoria in NT1 patient samples increasing 51% and 29% compared to controls, respectively.

In contrast, anti-HA and anti-NA antibodies against A/H1N1pre2009, A/H3N2, and B/Yamagata were not increased neither when comparing percentage of positive samples or GMT results, nor using multivariate logistic or linear regressions (Table 3). These findings were independent of HLA-DQB1*06:02 status, age, sex, flu season of collection. Most remarkable was the absence of changes in A/H3N2 antibody titers that was observed with both HAI and NAI assays, the latter assay being less sensitive to clade variations and antigen drift^21^.

### Validation of results on an extended sample set

To extend on these results, we tested a total of 441 samples including 181 NT1 patients versus 260 controls matched for sex and flu season of collection but were unable to perfectly match these by age (Table 2). Significantly increased percentage of positive samples for anti-HA antibodies against A/H1N1pdm2009(64.5% vs 28.7%) and B/Victoria (35.4% vs 27.1%) were still found in NT1 patients of this extended set. NT1 patients had 2.86 times and a 90 percent increased risk of infections by A/H1N1pdm2009, and B/Victoria respectively compared to controls. Further analysis on GMT of anti-NA antibodies revealed that in the extended sample set, NT1 patients also had increased anti-NA antibody level against A/H1N1pdm2009 (GMT: 722.2 vs 301.6) and B/Victoria (GMT: 415.4 vs 250.4)(Table 4). These results confirmed our data in a larger sample set spanning 25 years.

**Table 4.** Hemagglutinin and neuraminidase antibody levels in extended NT1 patients and controls.

| Subtypes/lineages | Hemagglutinin Inhibition Assay (HAI) |  |  |  | Neuraminidase Inhibition Assay (NAI) |  |  |  |
| --- | --- | --- | --- | --- | --- | --- | --- | --- |
|  | % of positive patients (N)* | % of positive controls (N)* | Odds Ratio(95%CI) | p-value# | GMT (95CI) of patients (N) | GMT (95CI) of controls (N) | β-coefficient (95%CI) | p-value# |
| Influenza A |  |  |  |  |  |  |  |  |
| All H1N1 | 64.6%(181) | 36.8% (258) | 2.20(1.43,3.39) | <0.01 | 180.4(132.1,246.3)(162) | 141.7(113.6,176.8)(246) | 0.08(-0.06,0.23) | 0.26 |
| H1N1pre2009 | 64.8% (88) | 51.1% (94) | 1.27(0.66,2.44) | 0.47 | 37.5(27.5,51.1)(76) | 41.8(32.5,53.8)(94) | 0.16(-0.03,0.35) | 0.11 |
| H1N1pdm2009 | 64.5% (93) | 28.7% (164) | 3.86(2.16,6.89) | <0.01 | 722.2(538.3,969.0)(86) | 301.6(233.0,390.3)(152) | 0.35(0.18,0.52) | <0.01 |
| H3N2 | 38.1% (181) | 31.4% (258) | 0.87(-0.56,1.36) | 0.55 | 54.4(45.4,65.3)(162) | 53.8(46.8,61.8)(246) | -0.03(-0.13,0.06) | 0.49 |
| Influenza B |  |  |  |  |  |  |  |  |
| B/Victoria | 35.4% (175) | 27.1% (258) | 1.90(1.21,2.97) | <0.01 | 415.4(342.6,503.7)(162) | 250.4(213.9,293.2)(246) | 0.22(0.11,0.32) | <0.01 |
| B/Yamagata | 14.9% (168) | 9.31% (247) | 1.17(0.62,2.23) | 0.63 | 233.7(200.9,271.8)(161) | 174.8(155.7,196.3)(243) | -0.01(-0.09,0.08) | 0.91 |

\*Using 40 as a cut-off value for being positive for HA antibody.
#Adjusted by age, sex, or other covariates when significant.

### Contributions of anti-HA and anti-NA antibodies against A/H1N1pdm2009 and B/Victoria to narcolepsy onset are independent

Correlations between anti-HA and anti-NA antibodies titers against the same strain were analyzed separately in patients and controls using Pearson correlation analysis. Correlations between anti-HA and anti-NA antibodies were found to be weak in both patients (r^2^=-0.04 to 0.25) (Table 5) and controls (r^2^=-0.10 to 0.34) (Table 5). This well-known phenomenon reflects surprisingly independent protection and immune reactions to HA and NA following flu infections^17^.

**Table 5.** Correlation analysis of HA and NA antibody titers on the same samples in NT1 patients and controls.

| Subtypes/lineages | NT1 patients |  |  |  | controls |  |  |  |
| --- | --- | --- | --- | --- | --- | --- | --- | --- |
|  | N | r | r<sup>2</sup> | *p value* | N | r | r<sup>2</sup> | *p value* |
| H1N1 | 162 | 0.06 | 0.003 | 0.48 | 246 | -0.10 | 0.01 | 0.12 |
| H1N1pre2009 | 76 | -0.04 | 0.002 | 0.77 | 94 | -0.03 | 0.001 | 0.79 |
| H1N1post2009 | 86 | 0.16 | 0.03 | 0.13 | 152 | 0.10 | 0.01 | 0.21 |
| H3N2 | 162 | 0.25 | 0.06 | <0.01 | 246 | 0.19 | 0.04 | <0.01 |
| B/Victoria | 162 | 0.18 | 0.03 | 0.02 | 246 | 0.34 | 0.12 | <0.01 |
| B/Yamagata | 161 | 0.02 | 0.0003 | 0.83 | 242 | 0.10 | 0.01 | 0.13 |

The lack of correlation between HAI and NAI assay results led us to explore how much these assays predict narcolepsy status. To do so, we conducted a full multivariate model (M₁), showing it significantly improved prediction of narcolepsy diagnosis over the age-only baseline (M₀) (likelihood-ratio test: χ^2^₄ = 50.98, p = 2.3 × 10^-10^). Global model performance increased substantially from M₀ to M₁ across all three metrics: McFadden’s pseudo-R^2^ rose from 0.029 to 0.192 (ΔR^2^ = 0.163), Tjur’s coefficient of discrimination from 0.037 to 0.236 (ΔD = 0.199), and AUC from 0.643 to 0.787 (ΔAUC = 0.144).

Multivariate analysis explored if anti-HA and anti-NA antibody associations were independent predictors of narcolepsy onset. Interestingly, anti-HA antibodies against A/H1N1pdm2009(β = 0.78, 95% CI [0.44, 1.12], p < 10^-6^) and anti-NA antibodies against B/Victoria (β = 0.69, 95% CI [0.31, 1.07], p < 10^-5^) were most strongly associated with narcolepsy. Anti-NA against A/H1N1pdm2009 showed a weaker but significant association (β = 0.42, 95% CI [0.07, 0.76], p = 0.02), while anti-HA against B/Victoria (β = 0.03, 95%CI[-0.28,0.34], p = 0.34) and age (β = −0.23, 95%CI[-0.57,0.11], p = 0.19) was not significant after inclusion of these other variables.

Shapley decomposition of each performance metric confirmed this ranking. Anti-HA antibodies against A/H1N1pdm2009 were the largest contributors (Shapley R^2^_McFadden = 0.078; Shapley D = 0.098; Shapley AUC = 0.081), closely followed by anti-NA against B/Victoria (Shapley R^2^_McFadden = 0.050; Shapley D = 0.056; Shapley AUC = 0.028). Anti-NA against A/H1N1pdm2009 contributed modestly (Shapley R^2^_McFadden = 0.031; Shapley D = 0.040; Shapley AUC = 0.034), and anti-HA against B/Victoria contributed negligibly to model performance (Shapley R^2^_McFadden = 0.004; Shapley D = 0.005; Shapley AUC = <0.001) (Table S2).

## Discussion

The search for an autoimmune basis for narcolepsy started in 1984 with the discovery of a strong association of narcolepsy with HLA-DR2 in Japan^22^ and subsequent identification of HLA-DQB1*06:02 as the causal association across all ethnic groups in 1992^23,24^. Involvement of an immune-related mechanism was subsequently confirmed by genome wide association studies [4], which outlined loci involved in antigen presentation to T-cell receptors and the possible engagement of CD4 and CD8 T cells, as well as association with INFAR1 and langerin which may reflect interactions with infectious agents based on functional analyses^4^. Increased T cell reactivity directed toward orexin has been reported, although the difference is not striking comparing with controls^5–7^. However, since then, autoantibodies have yet to be identified^2^.

Following these findings, efforts were made to understand triggers of narcolepsy onset. Initially, following observations that narcolepsy could follow strep throat infections, an association with Streptococcus Pyogenes was shown epidemiologically^25^ and by anti-streptolysin O antibody (ASO) studies^25^. However, it was not replicated in China^26^, suggesting either that S. Pyogenes is only associated with narcolepsy onset in some countries, or that differences exist in ASO reactivity following Streptococcus Pyogenes infections in China.

Follow up studies found that in children, who have abrupt onsets that can often be timed to the week, onset was seasonal and peaked in spring and summer^9^, further supporting the concept that upper airway infections in winter were important to onset of narcolepsy. Most strikingly, following the 2009 H1N1 pandemic, narcolepsy onsets increased ∼4.5 fold independent of vaccinations^9,27^, with a predicted delay of 4-5 months following the peak of incidence of infections. Similar increases were found in Europe^13^, the United States^8^, and Taiwan^28^, although not as preeminent.

In this study, we show that recent narcolepsy onsets are associated with increased antibody titers against some influenza subtypes/lineages (A/H1N1pdm2009 and B/Victoria), but not other (A/H3N2). This result, obtained over a 25-year period, suggests that not only A/H1N1pdm2009 but also B/Victoria may trigger narcolepsy on a specific genetic background that includes HLA-DQB1*06:02. The data is in line with other data suggesting increased narcolepsy onset not only following the 2009 H1N1 pandemic but also following influenza B/Victoria infections^13^. Taken together, these results leave little doubt regarding a major role for influenza in triggering narcolepsy. The fact that vaccination with Pandemrix® as opposed to an infection was a trigger of narcolepsy in 2010 indicates that it is the immune reaction, not the viral infection itself that mediates orexin-cell destruction.

Interestingly, influenza is a worldwide disease, which could also explain why the HLA association and genome wide architecture is universal across countries and cultures^2,4^, reflecting a common trigger. Study strength includes inclusion of NT1 subjects with samples collected within the same flu season preceding disease onset, and controls matched for age, sex, season, and year of collection. Comparisons performed otherwise would have been flawed, as strain specific infections depend on season and age, although this relation is complex^29^. Nonetheless, extending this dataset to a larger sample further away from onset with less well-matched controls confirmed this data, showing the finding to be remarkably robust.

The use of both HAI and NAI assays to confirm this finding is another strong aspect of our study. Among 10 proteins encoded by a typical flu genome HA and NA are the two main flu surface proteins of both flu A and flu B. For this reason, preexisting anti-HA and anti-NA antibodies are a first line of defense against exposure, and antibodies against either of these proteins have been shown to be independently associated with protection^30^, as reflected by the weak correlation we found in anti HA and anti NA antibody titers. Anti-HA antibody titers ≥40, are, however, generally used as indicative of protection, and indicate recent influenza infections or vaccinations^17^.

The importance of these two proteins is also reflected by their use in subtyping flu strains, with two main HA and NA polymorphic grouping defining H and N subtypes, notably currently circulating strains of A/H1N1 and A/H3N2 in humans. Many other A flu subtypes are present in animals, notably birds and swine, the main origin of human spillovers (typically the result of a reassortment of an animal virus with a human virus) resulting in regular pandemics^31^ with H5N1 being of current concern as this flu, of primary avian origin, has already spilled over to many mammalian species, with non-transmissible variants showing high lethality in exposed individuals.

Before A/H1N1pdm2009 pandemic domination as an H1N1 subtype, the main H1N1 subtype was a descendent of the 1918 Spanish flu pandemic, which had drifted significantly in the interim 90 years (this strain is noted as pre 2009H1N1 in this work)^31^. H3N2 is a descendent of the Asian flu of 1968, a pandemic that quickly followed the “Hong Kong” H2N2 pandemic of 1957, displacing this other virus^31^. Importantly, however, Influenza A/H3N2 has shown much greater antigenic drift than A/H1N1 leading to numerous circulating subclades^14^. This has resulted in more difficulties for the CDC in predicting which strain will circulate any given winter based on southern hemisphere circulation, this mismatch explaining frequent reduced efficacy of vaccination when this strain is dominant. The reason is that H3 hemagglutinin undergoes faster, more punctuated immune-driven evolution—particularly around key antigenic sites and glycosylation patterns—leading to more frequent escape from existing population immunity^14,32^. In this direction, it is notable that HA of H3N2 is generally more glycosylated than its counterpart in H1N1 and B/influenza, a difference of possible importance^33^. Influenza B, a related virus, does not reassort with influenza A and has a more distinct phylogeny. It shares 9 of 10 proteins, but has a different proton channel protein, BM2 instead of M2, which has a completely different phylogenic origin^34^. Until Covid 19 2020-2022, two main lineages of B flu, B/Victoria and B/Yamagata had been circulating, but B/Yamagata disappeared after this event^35^, as the number of flu infection plummeted secondary to containment procedures.

In this study, we measured HA and NA antibodies against pre 2009 and pdm2009H1N1, H3N2 and B/Victoria as well as B/Yamagata, strains known to have circulated during a large time interval of 25 years. Strain tested each year were matched to CDC strain advised for vaccination that year. As mentioned above, H3N2 is harder to predict and as a result more frequent H3N2 subclade mismatch have occurred with recommended strains. This well-known observation could have contributed to the lack of association with H3N2 antibodies found in this study. Nonetheless, this is rather unlikely to explain a completely negative association, as although both HA and NA show increased genetic drift in H3N2, it is discordant in NA and HA^21^ and would not likely result in a completely negative finding with both tests across 25 years as shown here. Rather, the fact A/H3N2 titers are not increased in either assay suggest this virus is not involved in triggering the disease. This is surprising, as A/H1N1 is phylogenetically much closer to A/H3N2 than B/Victoria^36^, the other virus with increased titers in recent onset narcolepsy.

Considering the findings that both influenza A/H1N1pdm2009 and B /Victoria antibody titers were increased in recent onset cases; one may wonder if a combination of multiple strains may not be needed in some cases. Analysis of our data rather does not support this hypothesis, as double positivity for A/H1N1 and influenza B was rare. It is also notable that increased antibody to both or either hemagglutinin and neuraminidase targets was found in recent onset cases and that titers across these assays were poorly correlated in both narcolepsy and controls. In fact, multivariate models predicting narcolepsy status, as overall goodness-of-fit measured by McFaden pseudo-R^2^ rose from 0.029 in the age-only model to 0.192 using both anti-HA and anti-NA antibodies against A/H1N1pdm2009 and B/Victoria. Finer analysis looking at individual contribution of each antibody while accounting for collinearity demonstrated that specifically anti-HA antibody against A/H1N1pdm2009 and anti-NA antibody against B/Victoria appeared as the dominant independent predictors of narcolepsy. This last result may suggest that it is neither reactivity to hemagglutinin nor neuraminidase that is involved in triggering narcolepsy, but rather immunity to another of the 10 proteins translated from influenza genomes. T cell cross reactivity between flu epitopes and orexin neurons, the likely cause of narcolepsy, would likely correlate with B cell responses to the same target, perhaps through epitope spreading^37^. In this direction, the PB1 polymerase stands out as an interesting candidate as it was of A(H1N1)pdm09 origin in Pandemrix^38^ and has the highest amount of homology among flu proteins across influenza A and B^39^.

What is the meaning of the higher titers observed here? In this study, we did not ask about recent history of flu vaccination or infection prior to onsets; it is thus unclear whether either or both could have been causal to increased titers, or if increased titer reflect a baseline difference independent of recent past exposure. In our study, most patients came from the US, where flu vaccination rate is 50% and vaccines are always unadjuvanted^40^. Considering that titers stay elevated at least 3-6 month following vaccination or infection^41^, the most likely explanation is a recent exposure to A/H1N1 or B/Victoria prior to narcolepsy onsets, perhaps causal to the autoimmune process. Considering all the data available, we hypothesize that risk following these influenza exposures is perhaps related to strength of the resulting immune reaction, which was strongest with AS03-aduvanted vaccines, versus infection, and is likely lowest following either non adjuvanted vaccination^42^ or more restricted in HA subunit specific vaccines^43^. This could also explain that even with Pandemrix®, unexplained variation in risk was observed across European countries^44^. This, plus the smaller effect of Arepanrix®, a similar AS03 adjuvanted vaccine used in Canada^45^ but later in the course of the pandemic and not first in children, has led to the hypothesis that the dramatic increase in Northern Europe was the result of both the vaccination plus the pandemic, which occurred concurrently there^11^. Not surprisingly, reports of post flu vaccination onset have been high since 2009 in pharmacovigilance databases^46^, thus only a carefully designed study would be able to establish any connection of narcolepsy risk with non-adjuvanted vaccines, which we believe will likely be small and perhaps even lower than post infection. It is also notable that infection often generates a broader immune response reflected in increasing both HA and NA antibodies for example, in comparison to vaccination^47^.

Another, not necessarily mutually exclusive explanation, could be that titers for these strains were elevated in at risk patients independent of recent infection or vaccination. This could be reflecting differential primoinfection with one of these strains or cumulative effects, not just the recent infection or vaccination. Repeated exposure by infection or vaccination often boosts titers to previously encountered strains and can broaden the antibody landscape, but boosting to the same strain produces smaller effects after repeated annual vaccination, especially when baseline titers are already high or antigenic imprinting is strong^48^. Studying patients further away from onset will be needed to evaluate if differences in titers are waning over time after onset as anticipated. Interestingly, flu antibody titers are known to mostly increase in the first 7 years of life^49^, with almost all children being infected with at least one flu strain by age 7. This is in line with the fact narcolepsy onset is extremely rare before 2 years old, and increases significantly around age 7, peaking in teens and early adulthood^50^. It is thus not unreasonable to speculate that in some of these cases the triggering event (including Pandemrix®) may have been a primoinfections with a specific strain, an event that could have lifelong consequences considering the original antigenic sin effects (the fact the first infection will determine future memory responses to common epitopes to other strains). The use of both assays also likely better reflect broad-based increases in immunity generally seen with infections rather than vaccinations.

## Conclusion

Although infectious diseases have long been suspected as triggers for autoimmunity, few associations have been definitively established: EBV for multiple sclerosis, Campylobacter Jejuni for Guillain Barre, and Streptococcus Pyogenes for Rheumatic Heart fever. In all these cases, however, the complexity of HLA associations and the larger size of the genome of the infectious agent may make it challenging to identify how these agents lead to autoimmunity. Our study reveals that both A/H1N1pdm2009 and B/Victoria flu strains are associated with narcolepsy, but not A/H3N2 or B/Yamagata. The specificity of the trigger and of the associated immunogenetics may facilitate identification of the mechanism linking infections with immunity in narcolepsy, with application to the understanding of other autoimmune diseases.

## Acknowledgments

This work was supported by grant NIH-5R01AI144798 to E.M and NIH-2U19AI057229 to M.D. NIH had no role in study design, data collection and analysis, decision to publish, or preparation of the manuscript. We thank all the subjects for their participation.

## Data availability

Anonymized data not published within this article will be made available by request from any qualified investigator.

