## Supplementary material for "Antibodies against influenza A/H1N1pdm2009 and B/Victoria strains but not A/H3N2 are increased in recent onset type 1 narcolepsy versus matched controls": Table S1, Table S2

This appendix has been provided by the authors to give readers additional information about the work.

SUPPLEMENTAL APPENDIX

List of Investigators

Han Yan, M.D., Ph.D.[1], Ling Lin, Ph.D.[1], Robin Guillard, Ph.D.[1], Jing Zhang, M.D.[1], Claudia Macaubas, Ph.D.[1], Fabio Pizza, M.D., Ph.D.[2,3], Francesco Biscarini,M.D.[2,3], Giuseppe Plazzi, M.D.[2,4], Vamsee Mallajosyula, Ph.D.[5], Mark Davis, Ph.D.[5,6], Holden Maecker, Ph.D.[6], Emmanuel Mignot, M.D., Ph.D.[1*]

[1]Center for Narcolepsy, Stanford University School of Medicine, Palo Alto, CA 94304; [2]IRCCS – Institute of Neurological Science, Bologna, 40139 Bologna, Italy; [3] Department of Biomedical and Neuromotor Sciences (DIBINEM), University of Bologna, Bologna, Italy; [4]Department of Biomedical, Metabolic and Neural Sciences, University of Modena and Reggio Emilia, 41125 Modena, Italy; [5]Institute of Immunity, Transplantation and Infection, Stanford University, Stanford, CA 94305, USA; [6]Department of Microbiology and Immunology, Stanford University School of Medicine, Stanford, CA, 94305, USA.

Table S1. Influenza strains used in HIA, and recombinant NA used in NIA

| Serotypes/lineages | Strain | #Catalog of recombinant NA |
|---|---|---|
| H1N1pre2009 | A/USSR/90/1977* | 40197-VNAHC |
| | A/Beijing/262/95 | |
| | A/New Caledonia/20/99 | |
| | A/Solomon Islands/3/2006 | |
| | A/Brisbane/59/2007 | |
| H1N1pdm2009 | A/California/07/2009* | 11058-VNAHC |
| | A/Michigan/45/2015* | 40568-V08B |
| | A/Victoria/2570/2019* | 40785-V08B |
| H3N2 | A/Sydney/5/97 | |
| | A/Panama/2007/99 | |
| | A/Fujian/411/2002 | |
| | A/California/07/2004 | |
| | A/Wisconsin/67/2005* | 40017-VNAHC |
| | A/Brisbane/10/2007 | |
| | A/Perth/16/2009 | |
| | A/Victoria/361/2011 | |
| | A/Texas/50/2012 | |
| | A/Switzerland/9715293/2013 | |
| | A/Hong Kong/4801/2014 | |
| | A/Singapore/INFIMH-16-0019/2016* | 40802-V08B |
| | A/Darwin/9/2021* | 40860-V08B |
| B/Victoria | B/Victoria/02/1987 | |
| | B/Shandong/7/97 | |
| | B/Hong Kong/330/2001 | |
| | B/Malaysia/2506/2004 | |
| | B/Brisbane/60/2008* | 40203-VNAHC |
| | B/Colorado/06/2017 | |
| | B/Washington/02/2019* | 40790-V08B |
| | B/Austria/1359417/2021* | 40863-V08B |
| B/Yamagata | B/Beijing/184/93 | |
| | B/Sichuan/379/99 | |
| | B/Shanghai/361/2002 | |
| | B/Florida/4/2006 | |
| | B/Wisconsin/1/2010 | |
| | B/Massachusetts/2/2012 | |
| | B/Phuket/3073/2013* | 40502-V07B |

*Strains of which recombinant NA were used for NAI assay.

Table S2.Results of multivariate logistic regression model

| Predictor | Beta | CI_lower | CI_upper | p_value | Shapley_R2_McFadden | Shapley_Tjur_D | Shapley_AUC |
|---|---|---|---|---|---|---|---|
| Age | -0.23 | -0.57 | 0.11 | 0.187 | | | |
| Anti-HA of A/H1N1pdm2009 | 0.78 | 0.44 | 1.12 | 0.001 | 0.078 | 0.098 | 0.081 |
| Anti-NA of A/H1N1pdm2009 | 0.42 | 0.07 | 0.76 | 0.019 | 0.031 | 0.04 | 0.034 |
| Anti-HA of B/Victoria | 0.03 | -0.28 | 0.34 | 0.856 | 0.004 | 0.005 | 0 |
| Anti-NA of B/Victoria | 0.69 | 0.31 | 1.07 | 0.001 | 0.05 | 0.056 | 0.028 |
